# Preserved metacognition despite impaired perception of intentionality cues in schizophrenia

**DOI:** 10.1101/2021.05.18.21257368

**Authors:** Ana Muthesius, Farina Grothey, Carter Cunningham, Susanne Hölzer, Kai Vogeley, Johannes Schultz

**Author notes:** To whom correspondence should be addressed. AM: Department of Psychiatry and Psychotherapy, University of Cologne, Kerpener Str. 62, 50937 Cologne, Germany. JS: Center for Economics and Neuroscience, University of Bonn, Nachtigallenweg 86, 53127 Bonn, Germany.

## Abstract

Social cognition and metacognition are frequently impaired in schizophrenia, and these impairments complicate recovery. Recent work suggests that different aspects of metacognition may not be impaired to the same degree. Furthermore, metacognition and the cognitive capacity being monitored need not be similarly impaired. Here, we assessed performance in detecting cues of intentional behaviour as well as metacognition about detecting those cues in schizophrenia. Thirty patients and controls categorized animations of moving dots into those displaying a dyadic interaction demonstrating a chase or no chase and indicated their confidence in these judgments. Perception and metacognition were assessed using signal detection theoretic measures, which were analysed using frequentist and Bayesian statistics. Patients showed a deficit compared to controls in detecting intentionality cues, but showed preserved metacognitive performance into this task. Our study reveals a selective deficit in the perception of intentionality cues, but preserved metacognitive insight into the validity of this perception. It thus appears that impairment of metacognition in schizophrenia varies across cognitive domains - metacognition should not be considered a monolithic stone that is either impaired or unimpaired.

**Highlights:**

- Impaired detection of chasing between two moving objects in schizophrenia
- No impairment in metacognition during this task in schizophrenia
- Preserved metacognition into social perception despite impaired social perception

## Introduction

Schizophrenia is one of the most disabling psychiatric diseases (Mathers et al., 2008). The poor functioning does not only depend on the characteristic psychotic symptoms, but also on deficits in cognitive functions (Arnon-Ribenfeld et al., 2017; Fett et al., 2011; Green, 1996). One type of cognitive function particularly important for understanding daily functioning in people with schizophrenia is social cognition (Fett et al., 2011; Green et al., 2008; Green and Leitman, 2008), which has been found to be impaired in numerous studies (Green et al., 2015; Savla et al., 2013). Social cognition can be studied for example by assessing the perception of social interactions presented in visual displays: Abstract, interacting moving objects can evoke complex human-like behaviours related to intentions such as chasing each other, wanting something or courting each other (Abell et al., 2000; Bassili, 1976; Blythe et al., 1999; Heider and Simmel, 1944; Santos et al., 2008). Perceiving such displays is associated with increased activity in a network of brain regions involved in social cognition (Blakemore et al., 2003; Castelli et al., 2000; Santos et al., 2010; Schultz et al., 2005). Patients with schizophrenia show impairments in correctly attributing intentions to interacting moving objects in different paradigms (Horan et al., 2009b; Langdon et al., 2017).

Functional outcomes in schizophrenia also crucially depend on the integrity of metacognitive awareness of one’
ss own cognitive capacities (Farrer and Franck, 2007; Frith and Done, 1988; Lysaker et al., 2013, 2011; Stephan et al., 2009). A growing number of studies investigated metacognition of perceptual processes. While a recent review has reported small-to-medium sized deficits in metacognition of perception (Rouy et al., 2021), several examples of preserved metacognition of perception exist. For example, one study reported impaired conscious yet preserved unconscious performance monitoring in a low-level visual perception task in schizophrenia (Charles et al., 2017). Another study reported that metacognitive performance monitoring was unrelated to psychosis when perceptual sensitivity was accounted for (Powers et al., 2017). Furthermore, self-reported schizotypy in healthy participants was found to contribute little to variations in metacognitive performance (Rouault et al., 2018). Preserved metacognitive abilities may allow to mitigate the impact of impaired cognitive skills through compensatory strategies (Arnon-Ribenfeld et al., 2017; Davies and Greenwood, 2018; Hasson-Ohayon et al., 2018; Koren et al., 2006; Lysaker et al., 2019, 2013, 2011). Therefore, a more differentiated investigation may be helpful for understanding self-monitoring deficits in schizophrenia.

Measuring metacognition is difficult as metacognition is not an explicit behaviour. One reliable and bias-free assessment method called meta-d’ (Maniscalco and Lau, 2014, 2012) is based on Signal Detection Theory (Green and Swets, 1966; Macmillan and Creelman, 1991). It consists in the combined assessment of discrimination between stimulus alternatives (perceptual performance) and reports of confidence in the discrimination responses. Accurate self-monitoring about perceptual judgments is reflected in higher confidence in true than in false percepts (metacognitive performance) (Fleming and Lau, 2014; Maniscalco and Lau, 2014, 2012). In addition, the meta-d’ method allows to measure metacognitive *efficiency*, a measure of self-monitoring adjusted for differences in perceptual performance (Maniscalco and Lau, 2012). This is particularly useful for distinguishing between cognitive and metacognitive skills in schizophrenia, where cognitive impairments are well known. In fact, the above-mentioned review reported inconclusive metacognitive deficits about perception in schizophrenia when controlling for perceptual performance (Rouy et al., 2021). Another aspect to consider is that metacognitive sensitivity is often higher for percepts about displays in which a target stimulus is present than for percepts about displays without stimulus (Fleming et al., 2010; Kanai et al., 2010). This may be due to the fact that more sensory evidence can be accumulated when reporting the presence rather than the absence of a target. For this reason, a variation of meta-d’ evaluates response-specific metacognitive sensitivity (Maniscalco and Lau, 2014).

To investigate metacognition in social perception, we asked schizophrenia patients and healthy controls to report perceived chasing between two interacting dots. This paradigm has revealed an association between perceived chasing and attribution of animacy in healthy individuals (Schultz et al., 2005) and a deficit in the perception of chasing in participants with autism (David et al., 2014). In addition, participants reported their confidence in their responses. We analysed the data using signal detection measures of performance and metacognition. Based on previous findings, we hypothesized that patients would perform worse than control persons when asked to judge the presence of chasing. Given mixed findings about perceptual metacognition, we were curious to assess metacognitive performance in this task.

## Methods

Thirty patients (age [mean ± SD] 33.6 ± 10.3 y, 8 females) satisfying DSM-V criteria for schizophrenia (DSM-V 295.90) as determined by consultation with treating psychiatrists and medical records and 30 healthy control subjects (age 34.1 ± 10.9 y, 8 females) individually matched for sex and age (±2 y), participated in this study. Participants’ age ranged from 19 to 60 years and were recruited for the present and another experiment, reported in a previous publication (Muthesius et al., 2020). All participants had normal or corrected-to-normal vision and were able to speak German sufficiently well to provide written informed consent, and to understand and follow the task instructions. The number of participants was determined by a power analysis optimised for the experiment reported in Muthesius et al. (2020). For this analysis, we assumed an effect size of 0.65 based on a prior study first reporting the task used (Schultz et al., 2019), an alpha error of P = .05 and a power of 80%. One patient did not complete the task and dropped out of the study. Patients were in- and outpatients recruited from the Department of Psychiatry at the University Hospital Cologne. Control participants were healthy volunteers recruited among the general population and showed no signs of pre-existent or current psychiatric conditions, as assessed in a clinical interview by a psychiatrist. Details of the sample are provided in Table 1. The study was carried out in compliance with the latest revision of the Declaration of Helsinki. All participants provided written informed consent. Ethical approval was provided by the University of Cologne Ethics Committee (18–265).

**Table 1:** Demographic Details and Assessment Instrument Scores of Participants.

| Measure | Patients | Controls |
| --- | --- | --- |
| Age (y) | 33.6 ± 10.3 | 34.1 ± 10.9 |
| Sex (N females; N males) | 8; 22 | 8; 22 |
| Disease duration (mo) | 106.9 ± 90.9 | N.A. |
| Medication (mg chlorpromazine/day equivalent) | 678.8 ± 416.1 | N.A. |
| PANSS total score | 50.6 ± 11.8 | N.A. |
| PANSS positive symptoms subscale | 11.73 ± 3.62 | N.A. |
| PANSS negative symptoms subscale | 14.13 ± 5.87 | N.A. |
| PANSS general symptoms subscale | 24.76 ± 5.55 | N.A. |
| CDSS | 0.07 ± 0.25 | N.A. |
| CGI | 4.07 ± 0.98 | N.A. |
| SOFAS | 56.53 ± 13.81 | N.A. |

*Note: CDSS, Calgary Depression Rating Scale for Schizophrenia; CGI, Clinical Global Impression-scale; PANSS, Positive and Negative Syndrome Scale for Schizophrenia; SOFAS, Social and Occupational Functioning Assessment Scale; N.A.: not applicable. Mean ± SD are shown except if otherwise stated.*

Patients’ illness duration varied between 0 (first-episode psychosis) and 25 years. Mean disease duration was 8.9 ± 7.6 (median: 6.5) years and widely varied in the sample: Fifteen patients ranged between 0 and 6 years, eight from 6 to 16 years, and seven from 16 to 25 years. Patients were excluded if diagnosed with any comorbid axis-I-disorder, significant medical illness including any past or present neurological disorder or acute substance intoxication, or if there was a risk of acute suicidality measured by item 8 of the Calgary Depression Rating Scale for Schizophrenia (CDSS) (Addington et al., 1993). We did not assess IQ but patients with an explicit diagnosis of mental retardation or learning disabilities were excluded. None of the participants were hospitalized coercively by order of the responsible local authorities. Patients’ symptoms severity were assessed using the Positive and Negative Syndrome Scale for Schizophrenia (PANSS) (Kay et al., 1987). All patients were under antipsychotic medication determined by the treating psychiatrist. Fourteen patients were treated with one, fourteen patients with two, and two patients with three antipsychotics simultaneously. Additionally, seven patients were treated with antidepressants, four with antiepileptics prescribed for anxiolysis and mood stabilization, and one with low potency neuroleptics. Chlorpromazine equivalent dosage of antipsychotic medication (**Table 1**) was calculated according to published conversion tables (Gardner et al., 2010; Leucht et al., 2014).

### Experimental Task

Participants performed a modified version of a social perception task (Schultz et al., 2005) and reported their confidence in their responses. In a two-alternative forced choice, participants reported whether two moving dots (one coloured red, the other blue; see **Figure 1A**) on a visual display chased each other or not. There were four types of trials in this 2 × 2 experimental design, resulting from the combination of two factors: interactive vs. control trials, and low or high level of cross-correlation between dot movements. The four types of trials contained different amounts of sensory evidence of chasing, as shown in **Figure 1B**. In interactive trials, the red dot followed the blue dot, and the dots thus chased each other. Control trials were based on the interactive trials, but modified such as to disrupt the chasing (the movements of the red dot were unchanged, but the movements of the blue dot were reversed in time and space compared to the interactive trials; see Schultz et al., 2005). Participants’ task was to discriminate interactive from control trials; their responses were considered correct if they responded “chasing” in interactive trials and if they responded “no chasing” in control trials.

**Figure 1.**
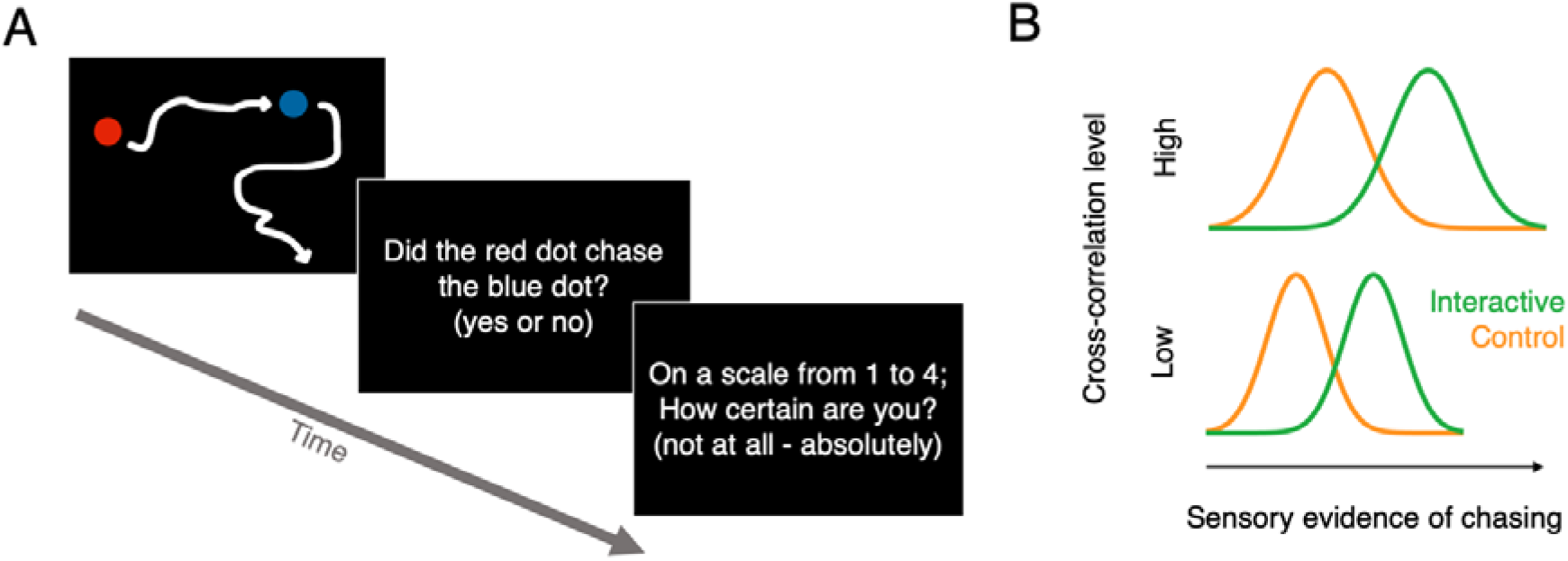
Stimuli, task and experimental design. **(A)** Example of the displays seen by the participants in the subsequent stages of each trial (stimulus; discrimination task; confidence rating). The white arrows represent the trajectories of the dots and were not shown on the screen. **(B)** The amount of sensory evidence of chasing varied both between interactive and control trials (the movement equation induced actual interactions between the moving dots in the interactive trials; these interactions were disrupted in the control trials) and as a function of the level of the cross-correlation level (more dot movements at high cross-correlation level increased actual chasing in interactive trials and induced apparent chasing in control trials). As a result, we expected participants to show a perceptual bias towards reporting chasing in trials with high cross-correlation level.

The level of cross-correlation is a parameter in a movement equation (detailed below) and determined the degree of influence that the dots had on each other’s position: with a high cross-correlation level, the dots chased each other more and also moved more than at the low cross-correlation level. In interactive trials with high cross-correlation, the dots really chased each other more than in interactive trials with low cross-correlation, because the degree of influence that the dots had on each other’s position was higher (see movement equation below). In control trials, the actual chasing was disrupted; however, randomly occurring similarities in the dot trajectories still occurred by chance. These randomly occurring trajectory similarities also influenced the percept of chasing, and were more likely to occur at the high cross-correlation level (for graded reports of perceived chasing at different cross-correlation levels, see Schultz et al., 2005). As a result, participants perceived more chasing in trials with high than with the low cross-correlation levels, in both control and interactive trials. In signal detection terms, observers would thus show a bias towards reporting chasing at the high cross-correlation level.

Our four trial types thus allowed us to assess and compare both discrimination capacity and perceptual bias in patients and control participants. A similar perceptual bias in patients and controls would indicate that patients processed the stimuli similarly to the controls and performed the task as desired. There were ten trials per trial type for a total of forty trials, presented in a randomized order. Each trial started with an animation sequence (4.3s) followed by participants’ chasing judgment and confidence rating [evaluated on a scale between 1 (= not confident at all) and 4 (= absolutely confident); self-paced].

The movement equation specified a time-series of positions for each object, where the new position of each object was determined by the previous position of both using a multivariate autoregressive process. The algorithm used the following equation to update the position of the dots [x(t+**Δ**t)] at each timepoint:

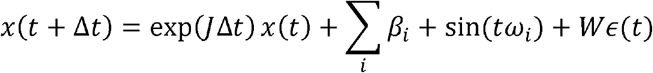

Here, *x*(*t*) are the coordinates of both objects, *J* is the system’s Jacobian controlling the dependencies:

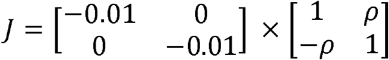

Δ*t* is the time step between two successive positions, ▯ = {1/7, 1/10, 1/2, 2/3} and ▯ = {1/100, 1/200, 1/50, 1/40} are terms driving the dot movements, *W* is a constant that scales the random term ∈(*t*) ∼ *N*(0,Δ*t*), and *ρ* is the cross-correlation parameter. Critically, the cross-correlation parameter determines the influence of each dot’s previous position on the other dot’s position: higher values make the red dot move closer to the blue dot and the blue dot move further away from the red dot; this therefore increases interactions between the dots. However, higher values of the cross-correlation parameter also increased the speed of both dots, leading to more motion (for a detailed analysis of the changes in dot speed and separation as a function of changes in cross-correlation level, see Schultz et al., 2005).

### Measurements

Perceptual performance was assessed through the signal detection theory measure d’, which quantified discrimination between trials with chasing and trials without chasing (higher values indicate better discrimination). Perceptual bias was assessed using the measure criterion, or bias, c’. Because a higher value of the cross-correlation parameter made the dots appear to chase each other more (in both the interactive and the control trials, leading to false positive chasing detection), we expected participants to tend to report chasing more frequently at the high cross-correlation level than at the low cross-correlation level, leading to a change in c’ between cross-correlation levels (lower values indicate a tendency to report chasing). The equations used to compute d’ and c’ are reported in the Supplementary materials.

We calculated three measures of metacognitive performance (Maniscalco and Lau, 2014, 2012) using the “meta-d’” toolbox (http://www.columbia.edu/~bsm2105/type2sdt/) (Maniscalco and Lau, 2012) implemented in Matlab (MATLAB R2021a, The Mathworks Inc., Natick, USA), with default settings. The first measure is the metacognitive sensitivity meta-d’, a response-bias free measure of how well a participant distinguishes between their correct and incorrect judgments. A participant who gives higher confidence ratings after correct judgments and lower confidence ratings after incorrect judgments has high metacognitive sensitivity. The second measure is the relative metacognitive sensitivity meta-d’/d’, also known as MRatio or metacognitive efficiency, which measures a participant’s metacognitive sensitivity given a certain level of task performance. The last measure is the response-specific metacognitive efficiency rs-meta-d’/d’, which involves calculating rs-meta-d’ separately for “yes” and “no” answers (Maniscalco and Lau, 2014). These measures were all estimated for each stimulus cross-correlation level and each participant using the maximum likelihood method, or a simpler sum of squares method in case the maximum likelihood fit did not converge to a solution.

### Statistical analysis

We performed two types of analyses. First, we analysed the dependent variables obtained in each trial of the task performed by our participants: chasing response (response to the task question “did the red dot chase the blue dot?”), response accuracy, confidence rating, and response time. These data were analysed using linear mixed regression models and linear mixed-effects logistic regression models, using Matlab 2021a using the function *fitglme*.*m*. All models contained an intercept, fixed effects (dummy variables coding the cross-correlation level and participant group, and for analyses of confidence and response time data, a dummy variable coding whether the response given was correct or not) and random effects (subject number). Models with interaction terms were compared to models without interaction terms using likelihood ratio tests implemented in Matlab’s function *compare*.*m*. Details of the analyses including the formulas used as well as detailed results are provided in the Supplementary materials.

Second, signal detection theory-based dependent variables were assessed using frequentist and Bayesian repeated-measures ANOVAs implemented in the software JASP, version 0.14.1 (www.jasp-stats.org), using default settings. Reported Bayes Factors (BF_10_) are odds in favour of the alternative hypothesis, i.e., ratios of the likelihood of the alternative hypothesis (=there is a difference between conditions) to the likelihood of the null hypothesis (=there is no difference between conditions), and are interpreted according to the currently recommended heuristic (Ly et al., 2016) based on Jeffrey’s rule (Jeffreys, 1961).

As meta-d’ and derived values are notoriously difficult to estimate from single-participant data with relatively low trial numbers, differences between participant groups can be directly estimated using a hierarchical Bayesian approach (Fleming, 2017). This approach enhances statistical power, incorporates uncertainty in group-level parameter estimates and avoids edge-correction confounds. We used the HMeta-d’ toolbox (https://github.com/metacoglab/HMeta-d) to estimate differences in meta-d’/d’ between patients and controls. This toolbox represents all participants’ data in a hierarchical graphical model, and uses a Markov chain Monte Carlo (MCMC) algorithm to estimate the joint posterior distribution of the model parameters, given the model specification and the data. In accordance with HMeta-d’ toolbox recommendations, early samples of the posterior distributions were discarded and three chains were run in order to diagnose convergence problems. Differences between participant groups were considered significant if the 95% highest-density intervals (HDI; the intervals containing 95% of the MCMC samples) of the posterior distribution of group-level difference parameters did not overlap with zero (Kruschke, 2014).

## Results

### Perceptual performance

#### Reports of chasing

First, we assessed whether changes in the stimuli led to the expected changes in perception of chasing in both patients and controls. We expected that participants would perceive chasing between the dots more often in displays in which there were interactions determined by the movement equation (interactive trials), and more often in trials with a higher cross-correlation level, irrespective of the presence of actual chasing (higher cross-correlation level induces a perceptual bias towards perceiving chasing; see Methods). Indeed, a mixed-effects logistic regression (see methods and Supplementary materials) revealed that participants reported chasing more often in the interactive trials (▯ = 1.54, SE = 0.07, 95% CI [1.41 1.67], t(2396) = 23.22, p << 0.001) and in trials with high cross-correlation level (▯ = 1.21, SE = 0.07, 95% CI [1.09 1.34], t(2396) = 18.57, p << 0.001; for full details of the model fit, see Supplementary materials). There was no significant difference between patients and control participants (▯ = 0.07, SE = 0.16, 95% CI [-0.24 0.38], t(2396) = 0.46, p = 0.65), nor interaction between interactive/control trials and cross-correlation level (▯ = -0.02, SE = 0.13, 95% CI [-0.24 0.28], t(2394) = 0.15, p = 0.88) or between cross-correlation level and participant group (▯= - 0.14, SE = 0.12, 95% CI [-0.38 0.11], t(2394) = -1.11, p = 0.27).

To better quantify the perceptual bias towards reporting chasing at higher cross-correlation level, we calculated the Signal Detection Theory measure of criterion or bias (c’, see Methods), expecting lower c’ values at higher cross-correlation level, which is indeed what we observed: c’ was lower in trials with high cross-correlation level than in those with low cross-correlation (**Figure 2A**; F(1,58) = 148.7, p << 0.001, ▯_p_^2^ = 0.72; two-way repeated measures ANOVA). There was no significant difference between patients and control persons (F(1,58) = 0.17, p = 0.68, ▯_p_^2^ < 0.01) nor interaction between participant group and cross-correlation level (F(1,58) = 0.37, p = 0.55, ▯_p_^2^ < 0.01). A Bayesian repeated-measures ANOVA revealed that the best model included only cross-correlation level (BF_M_ = 10.66). Post-hoc tests revealed extremely strong evidence of an effect of cross-correlation level (BF_10_ > 7.7*10^14^), and moderately strong evidence toward no difference between patients and controls (BF_10_ = 0.21). These data suggest that participants of both groups were similarly sensitive to stimulus manipulations.

**Figure 2.**
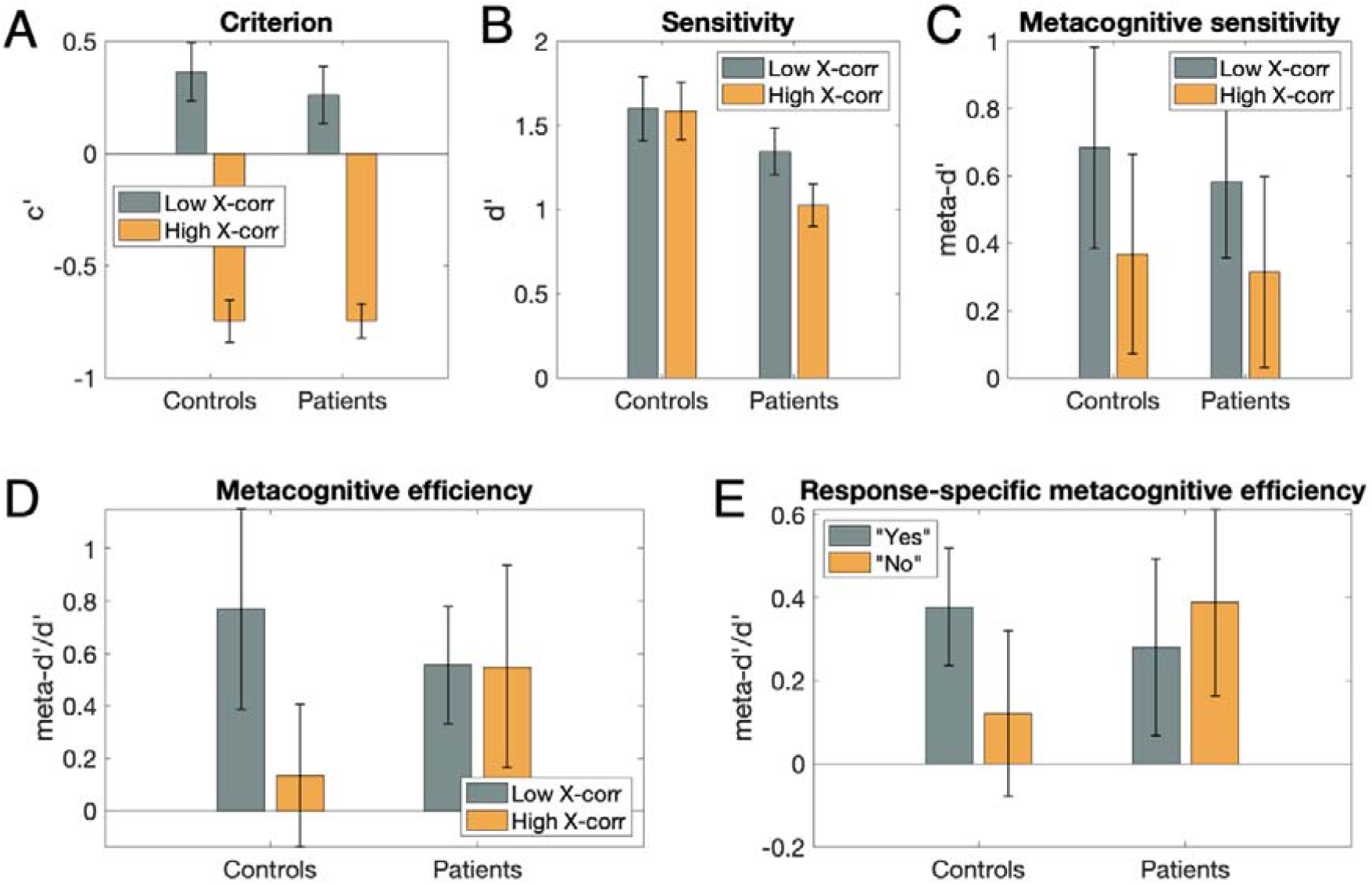
Results. **(A)** Criterion (c’) was lower in trials with high compared to low cross-correlation level in both participant groups, indicating a higher tendency to report chasing with high cross-correlation level. **(B)** Discrimination between chasing and no chasing (measured in d prime, d’) was reduced in patients compared to control persons. **(C-E)** No measure of metacognitive performance (C: meta-d’; D: meta-d’/d’; E: response-specific meta-d’) showed a significant difference between participant groups, trial types or response types. Error bars represent ±1 standard deviation, and boxes represent ±1 standard error of the mean.

#### Accuracy of reports of chasing

Next, we assessed participants’ accuracy at discriminating trials with actual chasing (interactive trials) from trials without actual chasing (control trials), as well as effects of the cross-correlation level on accuracy and differences in accuracy between patients and control participants. Average performance over all trials and participants was 72% correct. A mixed-effects regression model revealed that participants were better than chance (intercept: ▯ = 0.27, SE = 0.02, 95% CI [0.23 0.31], t(2397) = 14.14, p << 0.001), worse in high cross-correlation trials (▯ = -0.04, SE = 0.02, 95% CI [-0.07 -0.003], t(2397) = -2.16, p < 0.031), and patients were worse than controls (▯ = -0.06, SE = 0.02, 95% CI [-0.11 -0.016], t(2397) = - 2.64, p < 0.01). In the model with interaction term between cross-correlation level and participant group, this interaction was not significant (▯ = -0.12, SE = 0.11, 95% CI [-0.34 0.09], t(2396) = -1.13, p > 0.2). A mixed-effects logistic regression confirmed that participants made more errors in high cross-correlation level trials (▯ = -0.12, SE = 0.05, 95% CI [-0.23 -0.01], t(2397) = -2.17, p < 0.03), and that patients made more mistakes than control participants (▯ = -0.19, SE = 0.07, 95% CI [-0.33 -0.05], t(2397) = -2.60, p < 0.01). In the model with interaction term between cross-correlation level and participant group, this interaction was not significant (▯ = -0.12, SE = 0.11, 95% CI [-0.34 0.092], t(2396) = -1.13, p = 0.26). See **Supplementary Figure 1** for proportions of correct responses in all conditions.

We then assessed discrimination performance using the signal detection theory measure d’. Patients with schizophrenia were significantly less sensitive (i.e., had lower d’ values) in discriminating chasing from no chasing (**Figure 2B**; F(1,58) = 6.45, p = 0.01, ▯_p_^2^ = 0.10). There was no significant effect of cross-correlation level (F(1,58) = 1.19, p = 0.28, ▯_p_^2^ = 0.02) nor interaction between participant group and cross-correlation level (F(1,58) = 0.72, p = 0.40, ▯_p_^2^ = 0.01). A Bayesian repeated-measures ANOVA revealed that the best model included only participant group (BF_M_ = 3.43). Post-hoc tests revealed moderate evidence toward no effect of cross-correlation (BF_10_ = 0.25), and moderately strong evidence of a difference between patients and controls (BF_10_ = 3.25).

#### Confidence ratings

A mixed-effects regression model revealed that participants were more confident when they were correct (▯ = 0.24, SE = 0.04, 95% CI [0.16 0.33], t(2396) = 5.57, p << 0.001), and more confident in high cross-correlation trials (▯ = 0.17, SE = 0.04, 95% CI [0.09 0.25], t(2396) = 4.53, p << 0.001). Ratings from patients did not significantly differ from those of controls (▯ = -0.01, SE = 0.13, 95% CI [-0.26 0.25], t(2396) = -0.04, p > 0.9). A model with interaction terms did not reveal any significant interactions (see Supplementary materials). For distributions of confidence values in all conditions, see **Supplementary Figure 1**.

#### Response times

A mixed-effects regression model revealed that participants responded faster when they gave correct responses (▯ = -0.88, SE = 0.14, 95% CI [-1.16 -0.61], t(2396) = -6.42, p << 0.001), and that patients were slower than control participants (▯ = 0.73, SE = 0.33, 95% CI [0.08 1.38], t(2396) = 2.21, p < 0.027). The cross-correlation level did not significantly influence response times (▯ = -0.004, SE = 0.12, 95% CI [-0.24 0.23], t(2396) = 0.03, p > 0.9). A model with interaction terms did not reveal any significant interactions (see Supplementary materials). See **Supplementary Figure 1** for response times in all conditions.

#### Summary of perception findings

In sum, our data show that our task worked as desired: participants were better than chance at the task, and reported chasing more often in interactive trials. As we expected, stimuli created using a high cross-correlation level led to a perceptual bias towards reporting chasing in both patients and control participants. Crucially, patients were worse than control participants at the task, and showed reduced discrimination between interactive and control trials. Patients took longer to respond. Confidence ratings were higher when participants gave correct responses, but surprisingly these ratings were also higher in trials with high cross-correlation level, despite performance being lower in those trials.

### Metacognitive performance

#### Metacognitive sensitivity

Metacognitive sensitivity (meta-d’) did not differ significantly between patients and controls (**Figure 2C**; F(1,58) < 0.01, p = 0.98, ▯_p_^2^ < 0.001), did not vary with cross-correlation level (F(1,58) = 0.89, p = 0.35, ▯_p_^2^ = 0.015) and showed no interaction between cross-correlation level and participant groups (F(1,58) = 0.15, p = 0.70, ▯_p_^2^ = 0.003). A Bayesian repeated-measures ANOVA revealed that the null model best explained the data (BF_M_ = 5.83; including only the subject factor). Post-hoc tests revealed moderately strong evidence toward no effect of cross-correlation (BF_10_ = 0.22), and moderately strong evidence toward no difference between patients and controls (BF_10_ = 0.19).

#### Metacognitive efficiency

Metacognitive efficiency (meta-d’/d’) did also not differ between patients and controls (**Figure 2D**; F(1,58) = 0.61, p = 0.44, ▯_p_^2^ = 0.011), did not vary with cross-correlation level (F(1,58) = 0.50, p = 0.48, ▯_p_^2^ = 0.009) and showed no interaction between cross-correlation level and participant groups (F(1,58) = 1.71, p = 0.20, ▯_p_^2^ = 0.029). The hierarchical Bayesian analysis did not reveal evidence of differences between groups, neither for trials with low nor high cross-correlation (posterior distributions of difference estimates overlapped with 0: mean and 95% highest-density intervals of these distributions were 0.10; [-1.11, 1.21] and 0.61; [-0.53 1.71] for trials with low and high cross-correlation, respectively). See **Supplementary Figure 2** for differences in group posterior distributions of M-ratios obtained by hierarchical Bayesian estimation. These results suggest that there was no difference in metacognitive efficiency between patients and controls.

#### Response-specific metacognitive efficiency

Response-specific metacognitive efficiency (rs-meta-d’/d’) did not differ significantly between patients and controls (**Figure 2E**; F(1,49) = 0.49, p = 0.49, ▯_p_^2^ = 0.010), did not vary for “yes” vs. “no” responses (F(1,49) < 0.02, p = 0.90, ▯_p_^2^ < 0.001) nor with cross-correlation level (F(1,49) = 0.16, p = 0.69, ▯_p_^2^ = 0.003). Interactions between cross-correlation level and participant groups, cross-correlation level and response, and the three-way interaction were not significant (F(1,49) < 1.18, p > 0.28, ▯_p_^2^ < 0.023). There was a non-significant trend towards an interaction between response and participant groups (F(1,49) = 3.24, p = 0.078, ▯_p_^2^ = 0.062). Note that rs-meta-d’/d’ could not be estimated for 7 patients and 2 controls because of missing data for either yes or no responses in one or more stimulus conditions. A Bayesian repeated-measures ANOVA revealed that the null model best explained the data (BF_M_ = 24.10; including only the subject factor). Post-hoc tests revealed moderately strong evidence toward no difference between patients and controls and no effect of cross-correlation or response (BF_10_ values were respectively: 0.20; 0.11; 0.12).

#### Summary of metacognition findings

In sum, we found no evidence for differences in metacognitive performance between patients and controls in any measure of metacognition.

## Discussion

This study investigated perceptual and metacognitive performance in a social perception task in patients with schizophrenia and control persons. Consistent with established findings (Green et al., 2015; Green and Leitman, 2008), patients performed worse than control persons in detecting a social percept in displays of abstract social interactions. However, we found no significant differences in metacognitive performance between patients and controls. Bayesian tests indicated moderate effect sizes for both findings. In the context of recent occasional reports of preserved metacognitive capabilities (Charles et al., 2017; Powers et al., 2017; Rouy et al., 2021), our findings provide additional evidence for preserved metacognitive abilities.

Social perception in schizophrenia has been studied using a wide variety of tasks and measurements, ranging from descriptions of situations of daily life (Corrigan and Green, 1993; Toomey et al., 2002) to the interpretation of displays of interacting abstract objects (Horan et al., 2009b; Langdon et al., 2014, 2020; Roux et al., 2015). One advantage of displays of interacting abstract objects is that cues for intentional behaviour are solely carried by motion parameters, are independent from object appearance, and can be systematically varied in their quantity. This controlled approach to biological motion allows us to assess the sensitivity to cues about intentions-to-act inscribed in the presented movements irrespective of influences of the objects’ visual appearance or visual context. The psychophysical measures we used here allow a precise quantification of the sensitivity to cues of intentionality and do not depend for example on declarative verbal skills which can be disturbed in persons with schizophrenia.

Patients’ deficit in discriminating between chasing and non-chasing dot displays is unlikely to be due to basic stimulus processing or attention deficits, as we found similar effects of the cross-correlation parameter on perceptual biases in both patients and control persons. This suggests that patients could process the stimuli as we intended and were not simply overwhelmed by the task. The discrimination deficit indicates reduced sensitivity to visual cues for intentional interactions.

Several previous studies had investigated attributions of intentionality to displays of interacting agents in schizophrenia. One study described a deficit in the spontaneous attribution of social meaning to displays of interacting objects (Horan et al., 2009b). Participants verbally described each animation, and these descriptions were rated for the degree of intentionality attributed to the agents. Patients’ descriptions were rated lower in intentionality than those of control persons. This deficit was replicated in subsequent studies (Langdon et al., 2020, 2017). A chasing detection paradigm with several interacting objects developed by Gao and colleagues (2009) was used in three studies, with two studies reporting preserved detection of cues of intentionality (Langdon et al., 2020, 2014), while the third reported a deficit (Roux et al., 2015). Our present findings also reveal a deficit in chasing detection with a different display. In sum, a definitive consensus opinion about whether detection of intentionality cues in displays of interacting abstract agents is impaired in schizophrenia or not has not been reached yet.

We assessed metacognition about social perception by combining perceptual decisions (first-order task) and confidence ratings about these decisions. This procedure is considered to be the gold standard to investigate metacognition, because it allows to assess metacognition while taking into account impairments in first-order performance, which are frequent in schizophrenia: A recent meta-analysis found a global deficit of metacognition in schizophrenia driven by studies which did not equate first-order performance between groups, but no conclusive deficit among studies controlling for first-order performance (Rouy et al., 2021). We found no significant impairment in metacognitive performance, whether measured as meta d’, M-ratio or response-specific M-ratio. The data reported in **Figure 2D** show an unexpected, non-significant trend towards a reduced metacognitive efficiency in high cross-correlation trials in control participants, but no such trend in patients. This finding fits with the fact that in trials with high cross-correlation, participants reported higher confidence despite performing worse at the task. It seems that high cross-correlation trials induce a percept of chasing separately from actual, mathematically-determined interactions between the moving dots. We currently have no further explanation for this finding; to assess whether this trend is a real effect would require additional experiments, for example using a wider variation of stimuli. Our finding of the absence of significant impairment in metacognitive performance is compatible with the notion that not all types of metacognitive skills are similarly impaired in schizophrenia (see also (Rouy et al., 2021)). Other studies that have controlled for participant performance have also shown preserved components of metacognition in chronic schizophrenia, in the domains of episodic memory (Bacon and Izaute, 2009), detection of auditory signals (Powers et al., 2017), facial emotion recognition (Pinkham et al., 2018) and visual motion perception (Charles et al., 2017; Faivre et al., 2021). For example, Faivre and colleagues (Faivre et al., 2021) reported preserved metacognitive efficiency and sensitivity during perception of visual motion in people with chronic schizophrenia, and equivalent decisional mechanisms in patients and controls.

A more subtle investigation of metacognition consists in assessing the adaptability of metacognitive performance to the stimulus type, such as separate assessment of metacognitive performance for yes and no responses (Maniscalco and Lau, 2014). Indeed, confidence judgments about having perceived a target may be based on the amount of supporting evidence for the target. This is a somewhat different assessment than confidence judgments about not having perceived a target, which is based on the absence of evidence and might thus provide a poorer basis for discerning correct versus incorrect responses. A recent study reported that patients with schizophrenia do not show healthy controls’ adaptability of metacognitive performance to the type of visual motion stimuli (Koizumi et al., 2020). This could indicate that at least in some domains, patients with schizophrenia are atypical not in their metacognitive abilities, but rather in how their metacognitive capacity adapts to the demands of the cognitive function currently engaged. We found a non-significant trend toward a difference in response-specific metacognitive performance between patients and controls: M-ratios tended to be higher for yes-than for no-responses in control persons, as was observed in previous studies (Fleming et al., 2010; Kanai et al., 2010), and interestingly, this trend reversed in patients. While our findings did not reach significance, the adaptability of metacognitive performance to task demands in schizophrenia is a very interesting research topic that we might consider following up in subsequent studies. Assessing response-specific metacognitive performance is a sensitive approach that may allow us to better understand the mechanisms underlying the reported metacognitive difficulties previously reported in schizophrenia.

Some limitations of our study need to be acknowledged. First, our patient sample was small and heterogeneous: participants consisted of in- and outpatients, the sexes were not equally represented, and the duration of their disease varied widely. Second, while basic parameters such as age and sex were individually matched between patients and controls, we did not match the number of years of education and did not measure participants’ IQ. Lastly, our findings relate to processing of abstract stimuli in an experimental task, and thus our conclusions may not extend to real-life situations.

Social cognition and metacognition are challenging for people with schizophrenia, and are an important focus of psychotherapeutic interventions (Horan et al., 2009a; Moritz et al., 2014; Roberts et al., 2003; Wölwer et al., 2014). If preserved, specific metacognitive abilities can be used by patients to reconsider their interpretation of sensory signals they receive. For example, if patients learned to trust their metacognitive insight into a deficient perceptual process, they could learn to compensate or circumvent their deficit and improve the accuracy of their interpretations. Specifically, compensation could be achieved for example by taking more time and/or gathering more information before making a decision; the fact that patients took longer to respond in the present study supports the possibility that such a compensatory mechanism could be at play in the task we investigated here. Our results give some reason for optimism about this process regarding metacognition of social perception.

## Data Availability

The experiment and analysis code, and the data of the study are available here: https://osf.io/f8k9d/

## Acknowledgments

The authors have declared that there are no conflicts of interest in relation to the subject of this study. The experiment code, the data and code used for the analysis are posted on https://osf.io/f8k9d/.

## Author contributions

Ana Muthesius: conceptualization, investigation, data curation, writing – original draft, project administration. Farina Grothey: investigation, data curation. Carter Cunningham: software, formal analysis. Susanne Hölzer: investigation, data curation. Kai Vogeley: writing – review & editing. Johannes Schultz: conceptualization, data curation, software, formal analysis, writing – original draft, writing – review & editing, supervision.

## Supplementary methods

<u>Matlab code used to compute signal detection theory measures d’ and criterion c’ (=bias):</u>

~~~
% Calculate dprime for yes/no experiment, see Macmillan and Creelman 1991 p.8:
% Z-transform the hit (“chasing” responses in “interactive” trials) and fa-rates
(“chasing” responses in “control” trials):
hitZ = norminv(hitRate,0,1);
faZ = norminv(faRate,0,1);
% now subtract hitZ from faZ
dp = hitZ - faZ;
% Calculate criterion for yes/no experiment, see Macmillan and Creelman 1991
p.29:
c = -.5 * (hitZ + faZ);
~~~

## Supplementary results

**Supplementary Figure 1.**
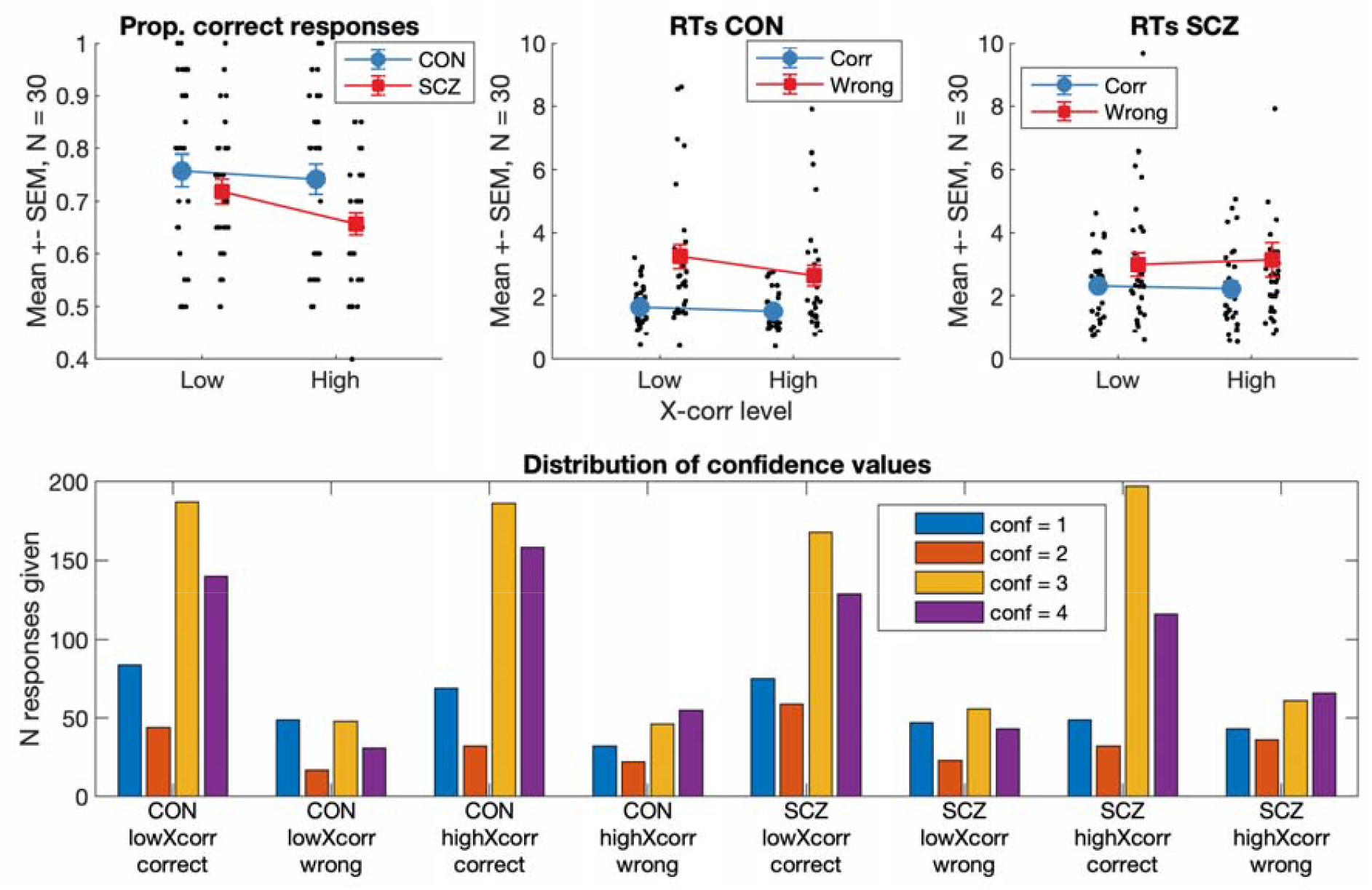
Top left: Average proportion correct per participant sorted by group (CON = controls, SCZ = patients) and stimulus type (low and high cross-correlation levels). Top middle and right: median response times in seconds for each participant sorted by group, stimulus type and response accuracy (Corr = correct response; Wrong = incorrect response). Bottom: distribution of confidence values per participant group, stimulus type, and response accuracy.

**Supplementary Figure 2.**
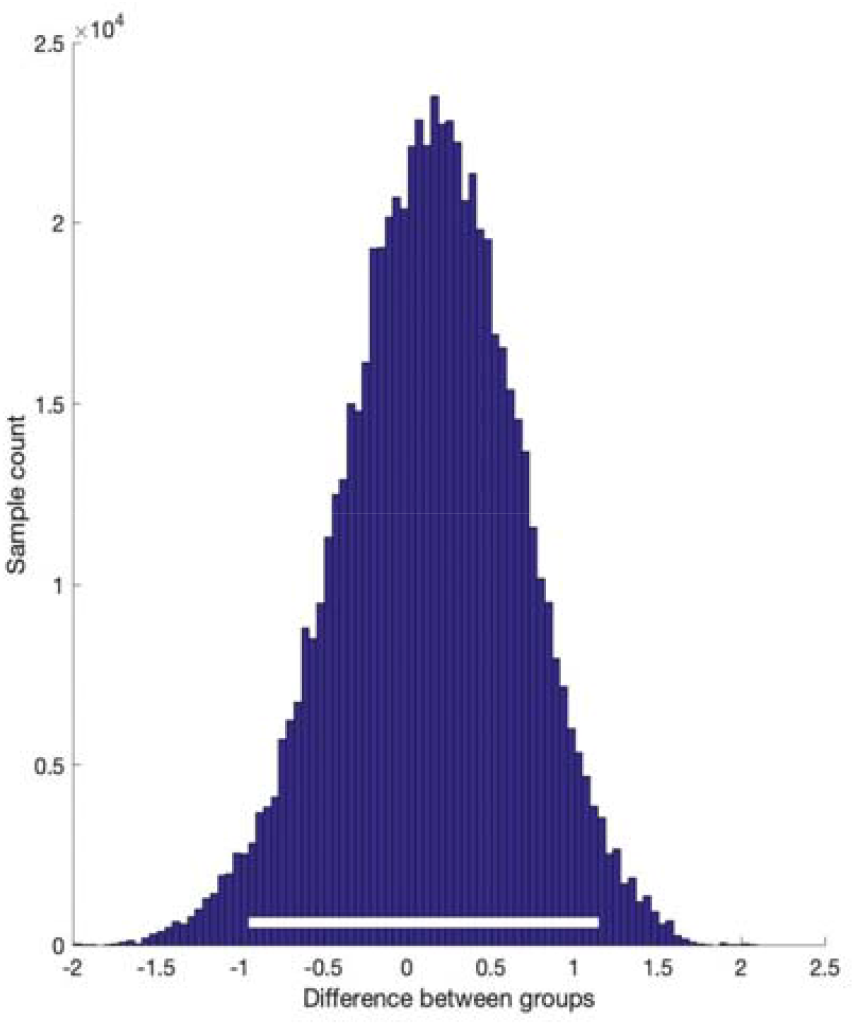
Difference in group posterior distributions of M-ratios obtained by hierarchical Bayesian estimation of metacognitive efficiency. Difference in group posteriors (in log units) between patients and controls, for all trials pooled across cross-correlation levels. Differences are shown in log units. The white bar indicates the 95% highest-density intervals. The bar overlaps zero, confirming the absence of a difference between patients and controls.

### Non-signal-detection-theory analyses of participant responses

#### Analysis of *chasing* responses

We ran a generalized linear mixed-effects logistic regression (probit) fitted by maximum likelihood using Laplace approximation on the *chasing* responses data (each trial of each participant, with values of 1 when the participant reported chasing, 0 when they did not). *interactive* was a dummy variable with values of 1 for interactive trials and 0 for control trials; *highXcorr* was a dummy variable with values of 1 for trials with high cross-correlation level and 0 for trials with low cross-correlation level; *patient* was a dummy variable with values of 1 for patient data and 0 for control participant data. In all analyses reported below, AIC is the model’s Akaike Information Criterion, BIC is the Bayesian Information Criterion, SE is the standard error, DF indicates the number of degrees of freedom, tStat is the effect’s t statistic, Lower and Upper are the 95% confidence intervals (CI) of the coefficient estimate, and significant fixed effects of interest are highlighted in **bold**:

##### Model information

|  |  |
| --- | --- |
| Number of observations | 2400 |
| Fixed effects coefficients | 3 |
| Random effects coefficients | 60 |
| Covariance parameters | 1 |
| Distribution | Binomial |
| Link | Probit |
| FitMethod | Laplace |

##### Formula

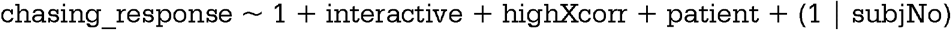

##### Model fit statistics

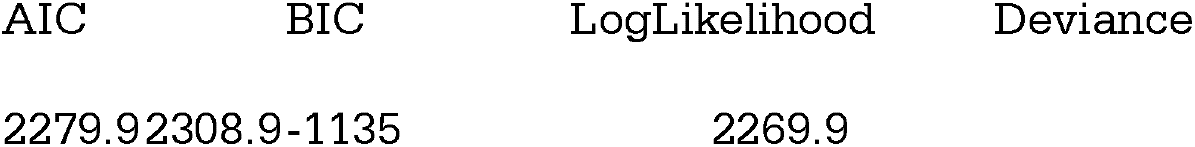

##### Fixed effects coefficients (95% CIs)

*Fixed effects coefficients (95% CIs):*
| *Name* | *Estimate* | *SE* | *tStat* | *DF* | *pValue* | *Lower* | *Upper* |
| --- | --- | --- | --- | --- | --- | --- | --- |
| (Intercept) | -1.15 | 0.12 | -9.44 | 2396 | 8.36e-21 | -1.39 | -0.91 |
| **interactive** | **1.54** | **0.07** | **23.22** | **2396** | **1.00e-107** | **1.41** | **1.67** |
| **highXcorr** | **1.21** | **0.07** | **18.57** | **2396** | **4.93e-72** | **1.09** | **1.34** |
| patient | 0.07 | 0.16 | 0.46 | 2396 | 0.65 | -0.24 | 0.38 |

##### Random effects covariance parameters

*Random effects covariance parameters:* Group: subject (60 Levels)
| *Name* | *Type* | *Estimate* |
| --- | --- | --- |
| (Intercept) | 'std' | 0.56 |

Model F statistic: F(3,2396) = 2013.3, p value = 0.0000

A comparison between the model without interaction (*NoInter*., see above) and the same model with interaction between *interactive* and *highXcorr* and between *highXcorr* and *patient* (*WithInter*.) was run using compare.m. LogLik is the log likelihood of each model, LRStat is the likelihood ratio test statistic. The results indicated that the model with interaction did not fit the data better:

##### Theoretical Likelihood Ratio Test

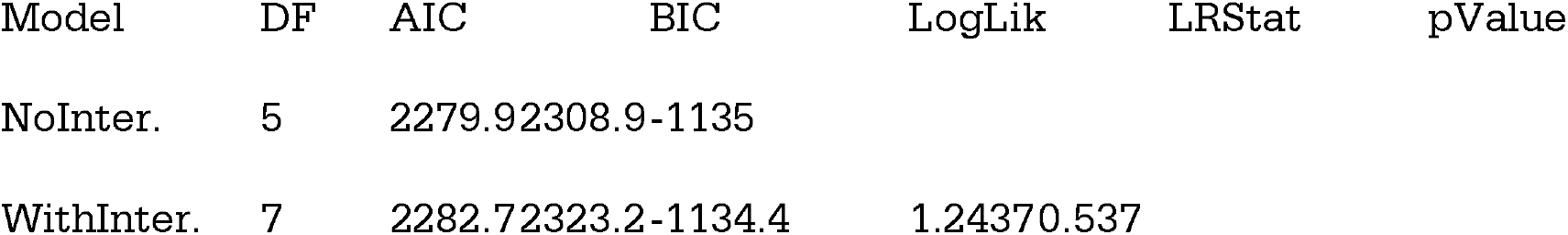

Note: the interaction terms in the model with interactions were not significant:

##### Fixed effects coefficients (95% CIs)

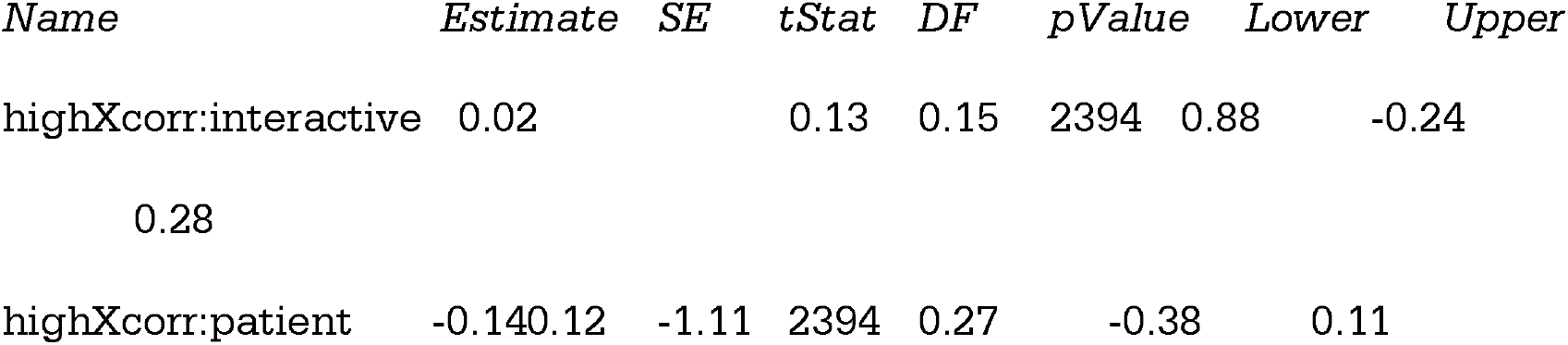

#### Analysis of accuracy: 1) comparison with chance

We ran a generalized linear mixed-effects regression fitted by maximum pseudo likelihood to the accuracy data. To compare accuracy with chance, we used as dependent variable *correctMinusChance*, which represented the responses given in each trial by each participant minus the response associated with chance (= 0.5). The dependent variable was thus 0.5 when the participant correctly reported chasing in interactive trials or no chasing in control trials, and -0.5 when their response was incorrect. The intercept thus assesses deviation from chance performance (0.5). Significant fixed effects of interest are highlighted in **bold**:

##### Model information

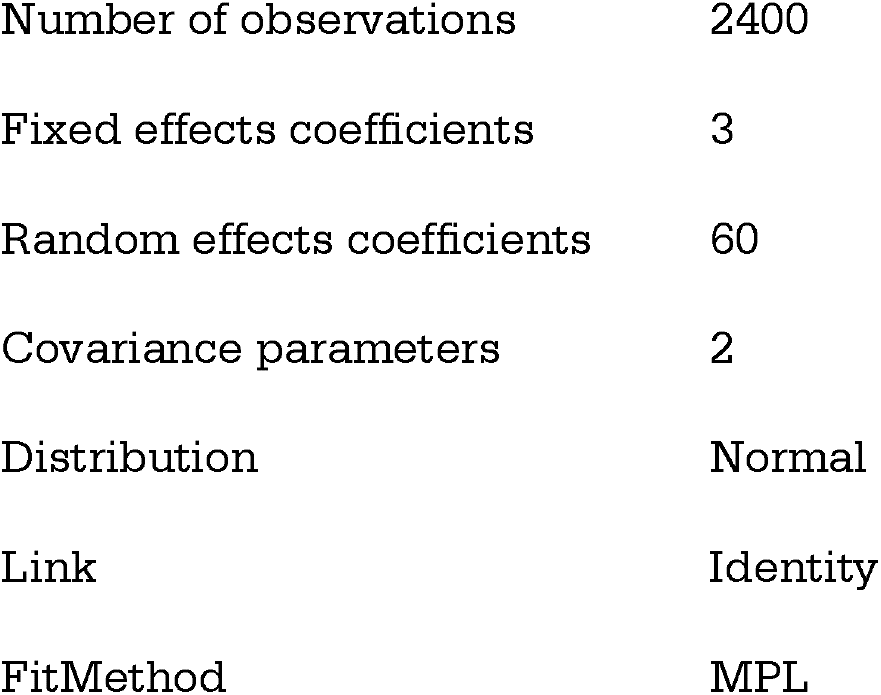

##### Formula

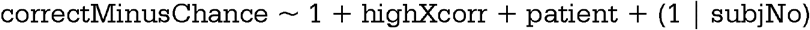

##### Model fit statistics

| AIC | BIC | LogLikelihood | Deviance |
| --- | --- | --- | --- |
| 2957.7 | 2986.6 | -1473.8 | 2947.7 |

##### Fixed effects coefficients (95% CIs)

*Fixed effects coefficients (95% CIs):*
| Name | Estimate | SE | tStat | DF | pValue | Lower | Upper |
| --- | --- | --- | --- | --- | --- | --- | --- |
| **(Intercept)** | **0.27** | **0.02** | **14.14** | **2397** | **1.11e-43** | **0.23** | **0.31** |
| **highXcorr** | **-0.04** | **0.02** | **-2.16** | **2397** | **0.031** | **-0.07** | **-0.003** |
| **patient** | **-0.06** | **0.02** | **-2.64** | **2397** | **0.008** | **-0.11** | **-0.016** |

##### Random effects covariance parameters

*Random effects covariance parameters:* *Group: subject (60 Levels)*
| Name | Type | Estimate |
| --- | --- | --- |
| *(Intercept)* | 'std' | 0.059162 |

Model F statistic: F(2,2397) = 17.3, p value = 0.0000

A comparison between the model without interaction (*NoInter*., see above) and the same model with interaction between *highXcorr* and *patient* (*WithInter*.) was run using compare.m and indicated that the model with interaction did not fit the data better:

##### Theoretical Likelihood Ratio Test

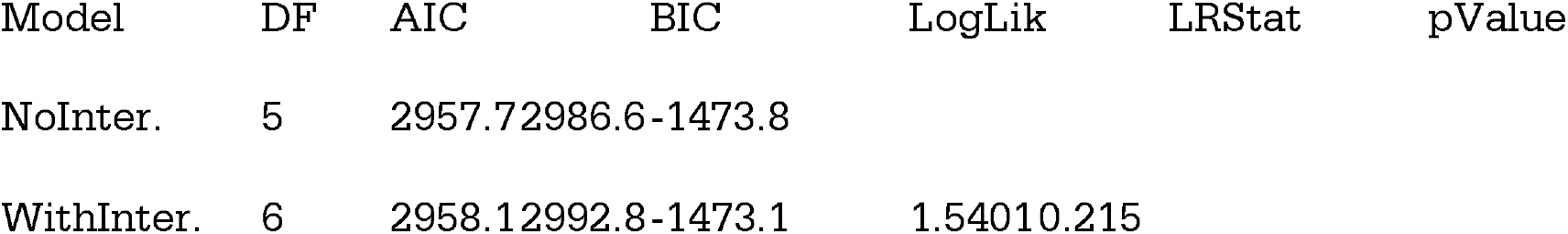

Note: the interaction term in the model with interaction was not significant:

##### Fixed effects coefficients (95% CIs)

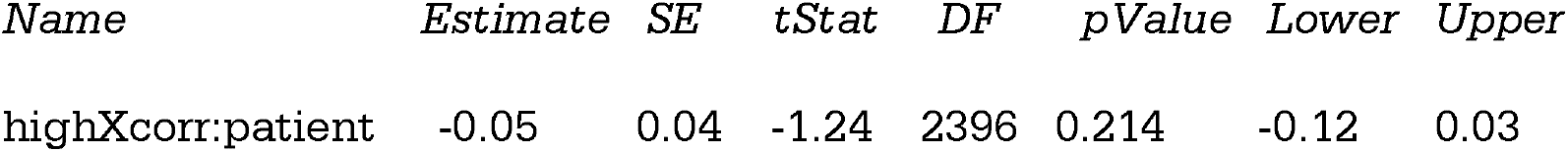

*Fixed effects coefficients (95% CIs):*
| Name | Estimate | SE | tStat | DF | pValue | Lower | Upper |
| --- | --- | --- | --- | --- | --- | --- | --- |
| highXcorr:patient | -0.05 | 0.04 | -1.24 | 2396 | 0.214 | -0.12 | 0.03 |

#### Analysis of accuracy: 2) logistic regression

As the simplest accuracy data were the values of 0 or 1 in each trial, we ran a generalized linear mixed-effects logistic regression (probit) fitted by maximum likelihood using Laplace approximation to the accuracy of the reported chasing responses. The dependent variable *correct_response* represented the responses given in each trial by each participant, with values of 1 when the participant correctly reported chasing in interactive trials or no chasing in control trials, and 0 when their response was incorrect. Significant fixed effects of interest are highlighted in **bold**:

##### Model information

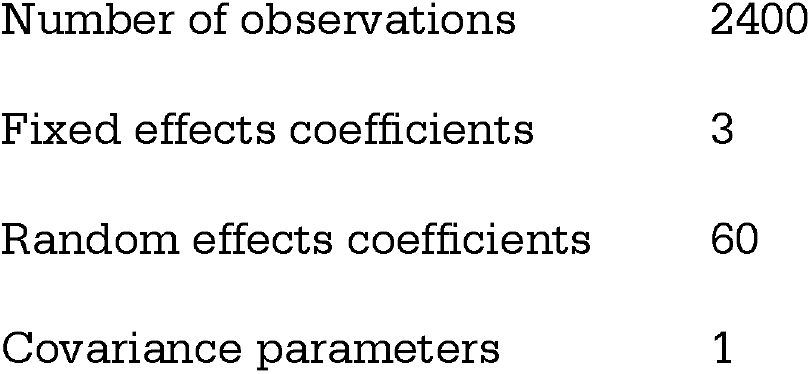

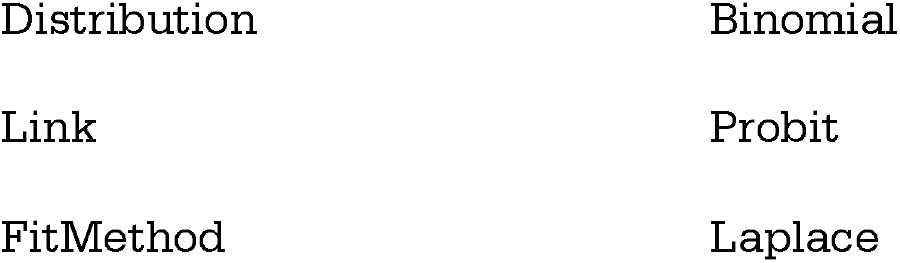

##### Formula

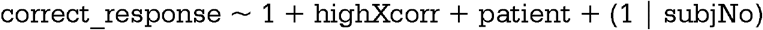

##### Model fit statistics

|  |  |  |  |
| --- | --- | --- | --- |
| AIC | BIC | LogLikelihood | Deviance |
| 2833.4 | 2856.5 | -1412.7 | 2825.4 |

##### Fixed effects coefficients (95% CIs)

*Fixed effects coefficients (95% CIs):*
| *Name* | *Estimate* | *SE* | *tStat* | *DF* | *pValue* | *Lower* | *Upper* |
| --- | --- | --- | --- | --- | --- | --- | --- |
| (Intercept) | 0.75 | 0.06 | 12.67 | 2397 | 1.23e-35 | 0.63 | 0.86 |
| **highXcorr** | **-0.12** | **0.05** | **-2.17** | **2397** | **0.029** | **-0.23** | **-0.01** |
| **patient** | **-0.19** | **0.07** | **-2.60** | **2397** | **0.009** | **-0.33** | **-0.05** |

##### Random effects covariance parameters

*Random effects covariance parameters:* Group: subject (60 Levels)
| *Name* | *Type* | *Estimate* |
| --- | --- | --- |
| (Intercept) | 'std' | 0.18 |

Model F statistic: F(2,2397) = 18.5, p value = 0.0000

A comparison between the model without interaction (*NoInter*., see above) and the same model with interaction between highXcorr and patient (*WithInter*.) was run using compare.m and indicated that the model with interaction did not fit the data better:

##### Theoretical Likelihood Ratio Test

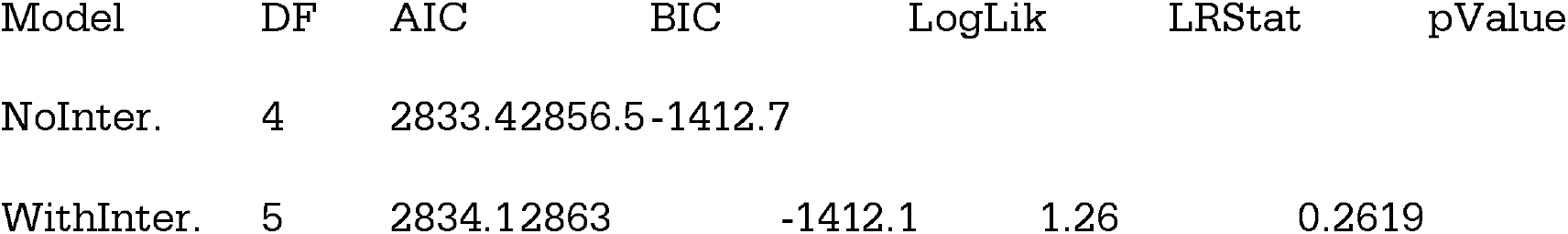

Note: the interaction term in the model with interaction was not significant:

##### Fixed effects coefficients (95% CIs)

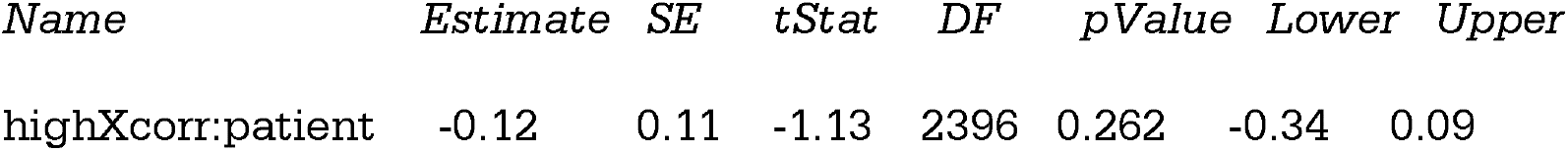

#### Analysis of confidence ratings

We ran a generalized linear mixed-effects regression fitted by maximum pseudo likelihood on the confidence rating data (each trial of each participant). Significant fixed effects of interest are highlighted in **bold**:

##### Model information

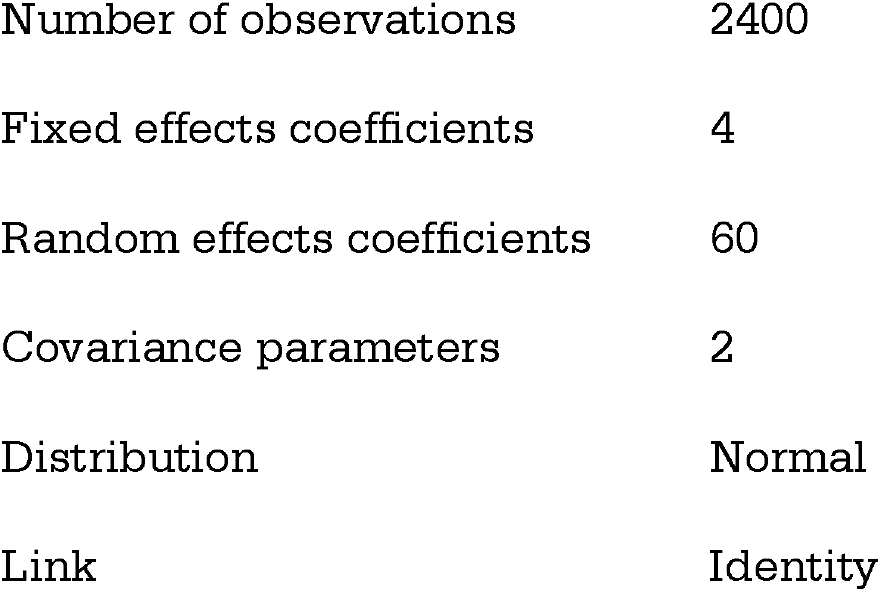
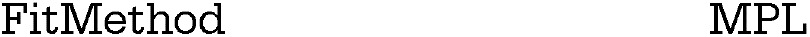

##### Formula

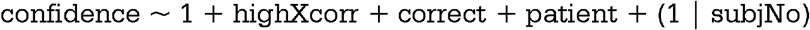

##### Model fit statistics

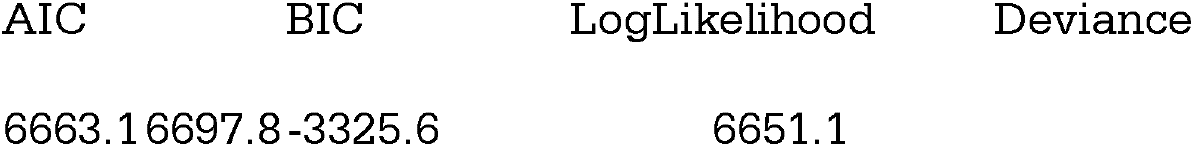

##### Fixed effects coefficients (95% CIs)

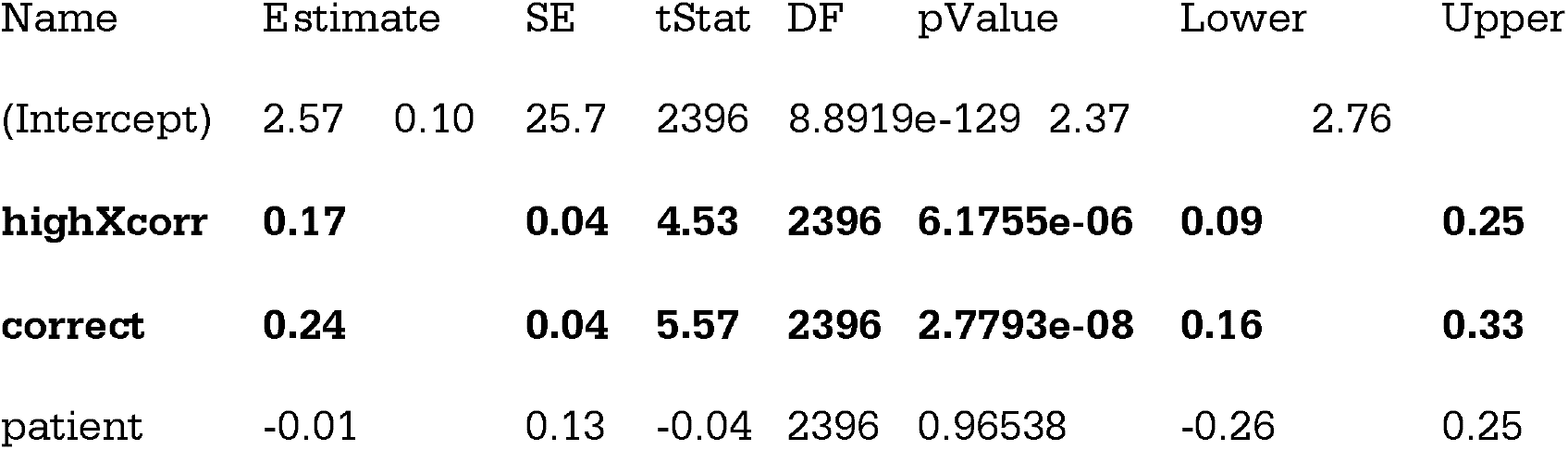

##### Random effects covariance parameters

*Random effects covariance parameters:* Group: subjNo (60 Levels)
| Name | Type | Estimate |
| --- | --- | --- |
| (Intercept) | std | 0.48201 |

Model F statistic: F(3,2396) = 216.7, p value = 0.0000

A comparison between the model without interaction (*NoInter*., see above) and the same model with interaction between correct and patient as well between correct and highXcorr (*WithInter*.) was run using compare.m and indicated that the model with interaction did not fit the data better:

##### Theoretical Likelihood Ratio Test

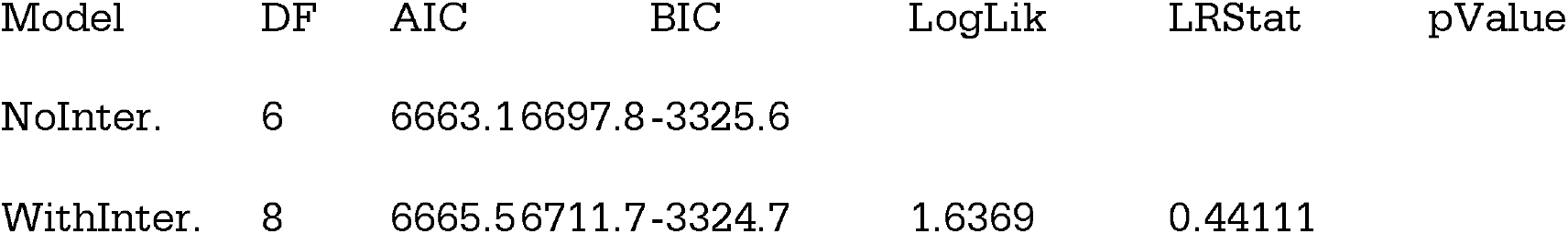

Note: the interaction terms in the model with interactions were not significant:

##### Fixed effects coefficients (95% CIs)

*Fixed effects coefficients (95% CIs):*
| Name | Estimate | SE | tStat | DF | pValue | Lower | Upper |
| --- | --- | --- | --- | --- | --- | --- | --- |
| highXcorr:correct | -0.10 | 0.09 | -1.18 | 2394 | 0.237 | -0.28 | 0.07 |
| highXcorr:patient | -0.04 | 0.08 | -0.57 | 2394 | 0.568 | -0.19 | 0.11 |

#### Analysis of response times data

We ran a generalized linear mixed-effects regression fitted by maximum pseudo likelihood on the response times data (RT, in seconds, data from each trial of each participant). *Correct* was a dummy variable with values of 0 for trials in which participants gave an incorrect response and values of 1 for trials in which participants gave a correct response. Significant fixed effects of interest are highlighted in **bold:**

##### Model information

|  |  |
| --- | --- |
| Number of observations | 2400 |
| Fixed effects coefficients | 4 |
| Random effects coefficients | 60 |
| Covariance parameters | 2 |
| Distribution | Normal |
| Link | Identity |
| FitMethod | MPL |

##### Formula

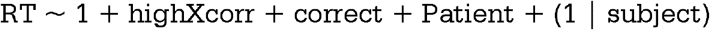

##### Model fit statistics

|  |  |  |  |
| --- | --- | --- | --- |
| AIC | BIC | LogLikelihood | Deviance |
| 12169 | 12204 | -6078.7 | 12157 |

##### Fixed effects coefficients (95% CIs)

*Fixed effects coefficients (95% CIs):*
| *Name* | *Estimate* | *SE* | *tStat* | *DF* | *pValue* | *Lower* | *Upper* |
| --- | --- | --- | --- | --- | --- | --- | --- |
| (Intercept) | 3.12 | 0.26 | 11.83 | 2396 | 1.9312e-31 | 2.60 | 3.64 |
| highXcorr | -0.004 | 0.12 | -0.03 | 2396 | 0.972 | -0.24 | 0.23 |
| **correct** | **-0.88** | **0.14** | **-6.42** | **2396** | **1.6869e-10** | **-1.16** | **-0.61** |
| **patient** | **0.73** | **0.33** | **2.21** | **2396** | **0.027** | **0.08** | **1.38** |

##### Random effects covariance parameters

*Random effects covariance parameters:* *Group: subject (60 Levels)*
| *Name* | *Type* | *Estimate* |
| --- | --- | --- |
| (Intercept) | 'std' | 1.191 |

Model F statistic: F(3,2396) = 144.7, p value = 0.0000

A comparison between the model without interaction (*NoInter*., see above) and the same model with interaction between highXcorr and patient and between correct and patient (*WithInter*.) was run using compare.m and indicated that the model with interaction did not fit the data better:

##### Theoretical Likelihood Ratio Test

| Model | DF | AIC | BIC | LogLik | LRStat | pValue |
| --- | --- | --- | --- | --- | --- | --- |
| NoInter. | 6 | 12169 | 12204 | -6078.7 |  |  |
| WithInter. | 8 | 12173 | 12219 | -6078.4 | 0.55306 | 0.75841 |

Note: the interaction terms in the model with interactions were not significant:

##### Fixed effects coefficients (95% CIs)

*Fixed effects coefficients (95% CIs):*
| Name | Estimate | SE | tStat | DF | pValue | Lower | Upper |
| --- | --- | --- | --- | --- | --- | --- | --- |
| highXcorr:correct | -0.18 | 0.28 | 0.28 | 2394 | 0.524 | -0.72 | 0.37 |
| highXcorr:patient | 0.08 | 0.24 | 0.39 | 2394 | 0.735 | -0.39 | 0.56 |

